# Detection of COVID-19 in smartphone-based breathing recordings using CNN-BiLSTM: a pre-screening deep learning tool

**DOI:** 10.1101/2021.09.18.21263775

**Authors:** Mohanad Alkhodari, Ahsan H. Khandoker

## Abstract

This study was sought to investigate the feasibility of using smartphone-based breathing sounds within a deep learning framework to discriminate between COVID-19, including asymptomatic, and healthy subjects. A total of 480 breathing sounds (240 shallow and 240 deep) were obtained from a publicly available database named Coswara. These sounds were recorded by 120 COVID-19 and 120 healthy subjects via a smartphone microphone through a website application. A deep learning framework was proposed herein the relies on hand-crafted features extracted from the original recordings and from the mel-frequency cepstral coefficients (MFCC) as well as deep-activated features learned by a combination of convolutional neural network and bi-directional long short-term memory units (CNN-BiLSTM). Analysis of the normal distribution of the combined MFCC values showed that COVID-19 subjects tended to have a distribution that is skewed more towards the right side of the zero mean (shallow: 0.59±1.74, deep: 0.65±4.35). In addition, the proposed deep learning approach had an overall discrimination accuracy of 94.58% and 92.08% using shallow and deep recordings, respectively. Furthermore, it detected COVID-19 subjects successfully with a maximum sensitivity of 94.21%, specificity of 94.96%, and area under the receiver operating characteristic (AUROC) curves of 0.90. Among the 120 COVID-19 participants, asymptomatic subjects (18 subjects) were successfully detected with 100.00% accuracy using shallow recordings and 88.89% using deep recordings. This study paves the way towards utilizing smartphone-based breathing sounds for the purpose of COVID-19 detection. The observations found in this study were promising to suggest deep learning and smartphone-based breathing sounds as an effective pre-screening tool for COVID-19 alongside the current reverse-transcription polymerase chain reaction (RT-PCR) assay. It can be considered as an early, rapid, easily distributed, time-efficient, and almost no-cost diagnosis technique complying with social distancing restrictions during COVID-19 pandemic.

## Introduction

Corona virus 2019 (COVID-19), which is a novel pathogen of the severe acute respiratory syndrome coronavirus 2 (SARS-Cov-2), appeared first in late November 2019 and ever since, it has caused a global epidemic problem by spreading all over the world [1]. According to the world heath organization (WHO) April 2021 report [2], there have been nearly 150 million confirmed cases and over 3 million deaths since the pandemic broke out in 2019. Additionally, the United States (US) have reported the highest number of cumulative cases and deaths with over 32.5 million and 500,000, respectively. These huge numbers have caused many healthcare services to be severely burdened especially with the ability of the virus to develop more genomic variants and spread more readily among people. India, which is one of the world’s biggest suppliers of vaccines, is now severely suffering from the pandemic after the explosion of cases due to a new variant of COVID-19. It has reached more than 17.5 million confirmed cases, setting it behind the US as the second worst hit country [2, 3].

COVID-19 patients usually range from being asymptomatic to developing pneumonia and in severe cases, death. In most reported cases, the virus remains incubation for a period of 1 to 14 days before the symptoms of an infection start arising [4]. Patients carrying COVID-19 have exhibited common signs and symptoms including cough, shortness of breath, fever, fatigue, and other acute respiratory distress syndromes (ARDS) [5, 6]. Most infected people suffer from mild to moderate viral symptoms, however, they end up by being recovered. On the other hand, patients who develop severe symptoms such as severe pneumonia are mostly people over 60 years of age with conditions such as diabetes, cardiovascular diseases (CVD), hypertension, and cancer [4, 5]. On most cases, the early diagnosis of COVID-19 helps in preventing its spreading and development to severe infection stages. This is usually done by following steps of early patient isolation and contact tracing. Furthermore, timely medication and efficient treatment reduces symptoms and results in lowering the mortality rate of this pandemic [7].

The current gold standard in diagnosing COVID-19 is the reverse-transcription polymerase chain reaction (RT-PCR) assay [8,9]. It is the most commonly used technique worldwide to successfully confirm the existence of this viral infection. Additionally, examinations of the ribonucleic acid (RNA) in patients carrying the virus provide further information about the infection, however, it requires longer time for diagnosis and is not considered as accurate as other diagnostic techniques [10]. The integration of computed tomography (CT) screening is another effective diagnostic tool (sensitivity ≥ 90%) that often provides supplemental information about the severity and progression of COVID-19 in lungs [11, 12]. CT imaging is not recommended for patients at the early stages of the infection, i.e., showing asymptomatic to mild symptoms. It provides useful details about the lungs in patients with moderate to severe stages due to the disturbance in the pulmonary tissues and its corresponding functions [13]. However, CT imaging may not be available in all public healthcare services, especially for countries who are swamped with the pandemic, due to its costs and additional maintenance requirements. Therefore, biological signals, such as coughing and breathing sounds, could be another promising tool to indicate the existence of the viral infection [14]. In addition, due to the simplicity in recording respiratory signals, lung sounds could carry useful information about the viral infection, and thus, could set an early alert to the patient before moving on with further medication procedures. In addition, the new emerging algorithms in artificial intelligence (AI) could be a key to enhance the sensitivity of detection for positive cases due to its ability to generalize over a wide set of data [15].

Many studies have investigated the information carried by respiratory sounds in patients tested positive for COVID-19 [16–18]. Furthermore, it has been found that vocal patterns extracted from COVID-19 patients’ speech recordings carry indicative biomarkers for the existence of the viral infection [19]. In addition, a telemedicine approach was also explored to observe evidences on the sequential changes in respiratory sounds as a result of COVID-19 infection [20]. Most recently, AI was utilized in one study to recognize COVID-19 in cough signals [21] and in another to evaluate the severity of patients’ illness, sleep quality, fatigue, and anxiety through speech recordings [22].

Despite of the high levels of performance achieved in the aforementioned AI-based studies, further investigations on the capability of respiratory sounds in carrying useful information about COVID-19 are still required, especially when embedded within the framework of sophisticated AI-based algorithms. Furthermore, due to the explosion in the number of confirmed positive COVID-19 cases all over the world, it is essential to ensure providing a system capable of recognizing the disease in signals recording through portable devices, such as computers or smartphones, instead of regular clinic-based electronic stethoscopes.

Motivated by the aforementioned, a complete deep learning approach is proposed in this paper for a successful detection of COVID-19 using only breathing sounds recorded through a microphone of a smartphone device (Fig. 1). The proposed approach serves as a rapid, no-cost, and easily distributed pre-screening tool for COVID-19, especially for countries who are in a complete lockdown due to the wide spread of the pandemic. Although the current gold standard, RT-PCR, provides high success rates in detecting the viral infection, it has various limitations including the high expenses involved with equipment and chemical agents, requirement of expert nurses and doctors for diagnosis, violation of social distancing, and the long testing time required to obtain results (2-3 days). Thus, the development of a deep learning model overcomes most of these limitations and allows for a better revival in the healthcare and economic sectors in several countries.

**Fig 1.**
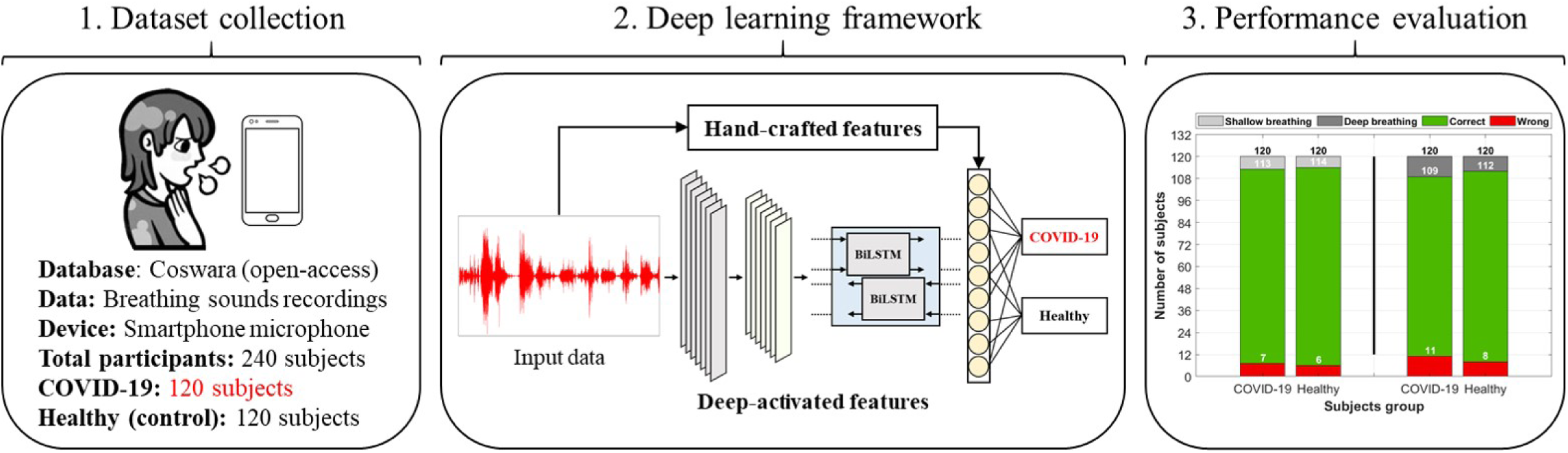
A graphical abstract of the complete procedure followed in this study. The input data includes breathing sounds collected from an open-access database for respiratory sounds (Coswara [23]) recorded via smartphone microphone. The data includes a total of 240 participants, out of which 120 subjects were suffering from COVID-19, while the remaining 120 were healthy (control group). A deep learning framework was then utilized based on hand-crafted features extracted by feature engineering techniques, as well as deep-activated features extracted by a combination of convolutional and recurrent neural network. The performance was then evaluated and further discussed on the use of artificial intelligence (AI) as a successful pre-screening tool for COVID-19.

Furthermore, the novelty of this work lies in utilizing smartphone-based breathing recordings within this deep learning model, which, when compared to conventional respiratory auscultation devices, i.e., electronic stethoscopes, are more preferable due to their higher accessibility by wider population. This plays an important factor in obtaining medical information about COVID-19 patients in a timely manner while at the same time maintaining an isolated behaviour between people. Additionally, this study covers patients who are mostly from India, which is severely suffering from a new genomic variant (first reported in December 2020) of COVID-19 capable of escaping the immune system and most of the available vaccines [2, 24]. Thus, it gives an insight on the ability of AI algorithms in detecting this viral infection in patients carrying this new variant, including asymptomatic. Lastly, the study presented herein investigates signal characteristics contaminated within shallow and deep breathing sounds of COVID-19 and healthy subjects through deep-activated attributes (neural network activations) of the original signals as well as wide attributes (hand-crafted features) of the signals and their corresponding mel-frequency cepstrum (MFC). The utilization of one-dimensional (1D) signals within a successful deep learning framework allows for a simple, yet effective, AI design that does not require heavy memory requirements. This serves as a suitable solution for further development of telemedicine and smartphone applications for COVID-19 (or other pandemics) that can provide real-time results and communications between patients and clinicians in an efficient and timely manner. Therefore, as a pre-screening tool for COVID-19, this allows for a better and faster isolation and contact tracing than currently available techniques.

## Materials and Methods

### Dataset collection and subjects information

The dataset used in this study was obtained from Coswara [23], which is a project aiming towards providing an open-access database for respiratory sounds of healthy and unhealthy individuals, including those suffering from COVID-19. The project is a worldwide respiratory data collection effort that was first initiated in August, 7th 2020. Ever since, it has collected data from more than 1,600 participants (Male: 1185, Female: 415) from allover the world (mostly Indian population). The database was approved by the Indian institute of science (IISc), human ethics committee, Bangalore, India, and conforms to the ethical principles outlined in the declaration of Helsinki. No personally identifiable information about participants was collected and the participants’ data was fully anonymized during storage in the database.

The database includes breath, cough, and voice sounds acquired via crowdsourcing using an interactive website application that was built for smartphone devices [25]. The average interaction time with the application was 5-7 minutes. All sounds were recorded using the microphone of a smartphone and sampled with a sampling frequency of 48 kHz. The participants had the freedom to select any device for recording their respiratory sounds, which reduces device-specific bias in the data. The audio samples (stored in .WAV format) for all participants were manually curated through a web interface that allows multiple annotators to go through each audio file and verify the quality as well as the correctness of labeling. All participants were requested to keep a 10 cm distance between the face and the device before starting the recording.

So far, the database had a COVID-19 participants’ count of 120, which is almost 1-10 ratio to healthy (control) participants. In this study, all COVID-19 participants’ data was used, and the same number of samples from the control participants’ data was randomly selected to ensure a balanced dataset. Therefore, the dataset used in this study had a total of 240 subjects (COVID-19: 120, Control: 120). The demographic and clinical information of the selected subjects is provided in Table 1. Furthermore, only breathing sounds of two types, namely shallow and deep, were obtained from every subject and used for further analysis (examples from the shallow breathing dataset are shown in Fig. 2). To ensure the inclusion of maximum information from each breathing recording as well as to cover at least 2-4 breathing cycles (inhale and exhale), a total of 16 seconds were considered, as the normal breathing pattern in adults ranges between 12 to 18 breaths per minute [26]. All recordings with less than 16 seconds were padded with zeros. Furthermore, the final signals were resampled with a sampling frequency of 4 kHz.

**Table 1.**
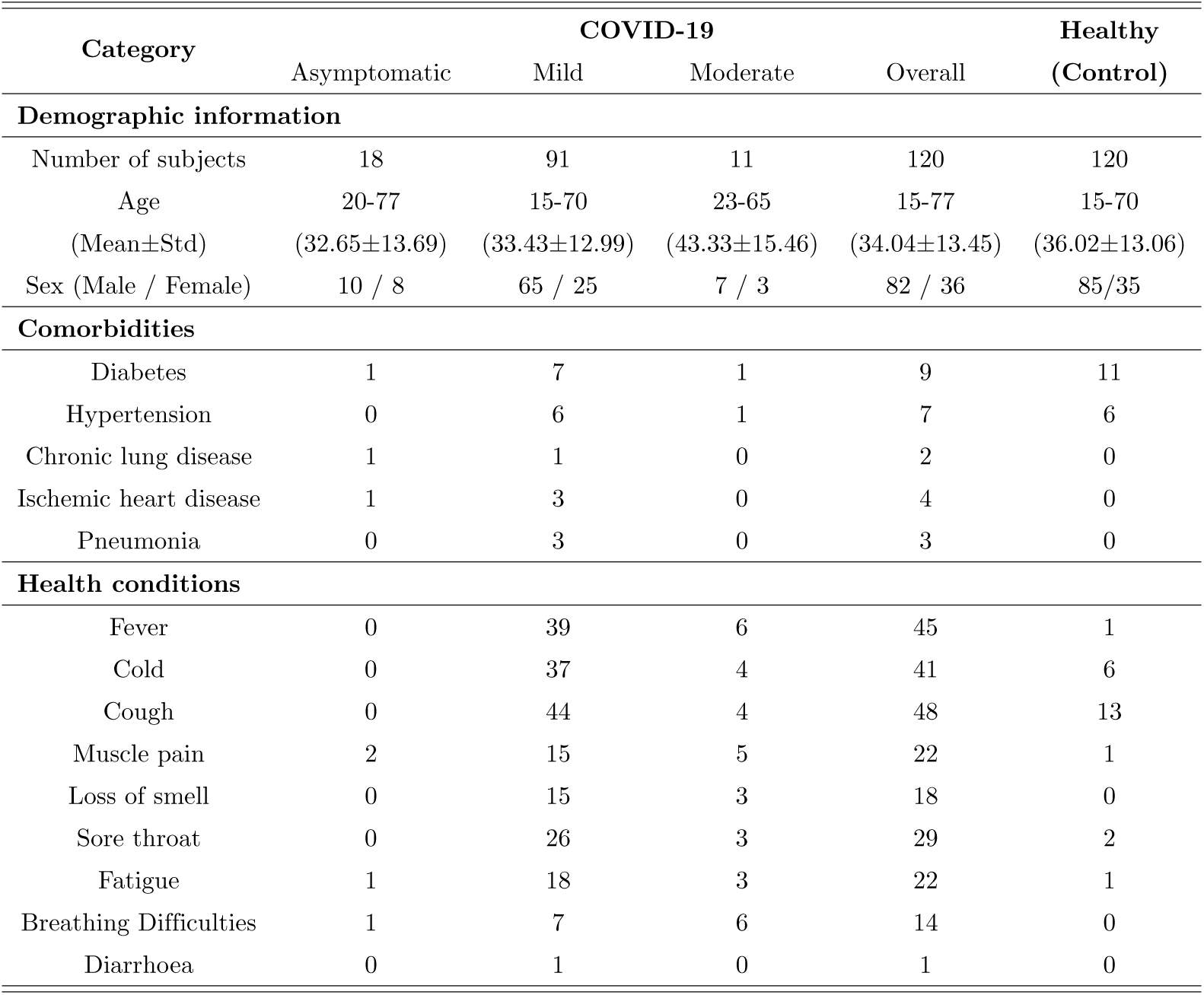
The demographic and clinical information of COVID-19 and healthy (control) subjects included in the study.

**Fig 2.**
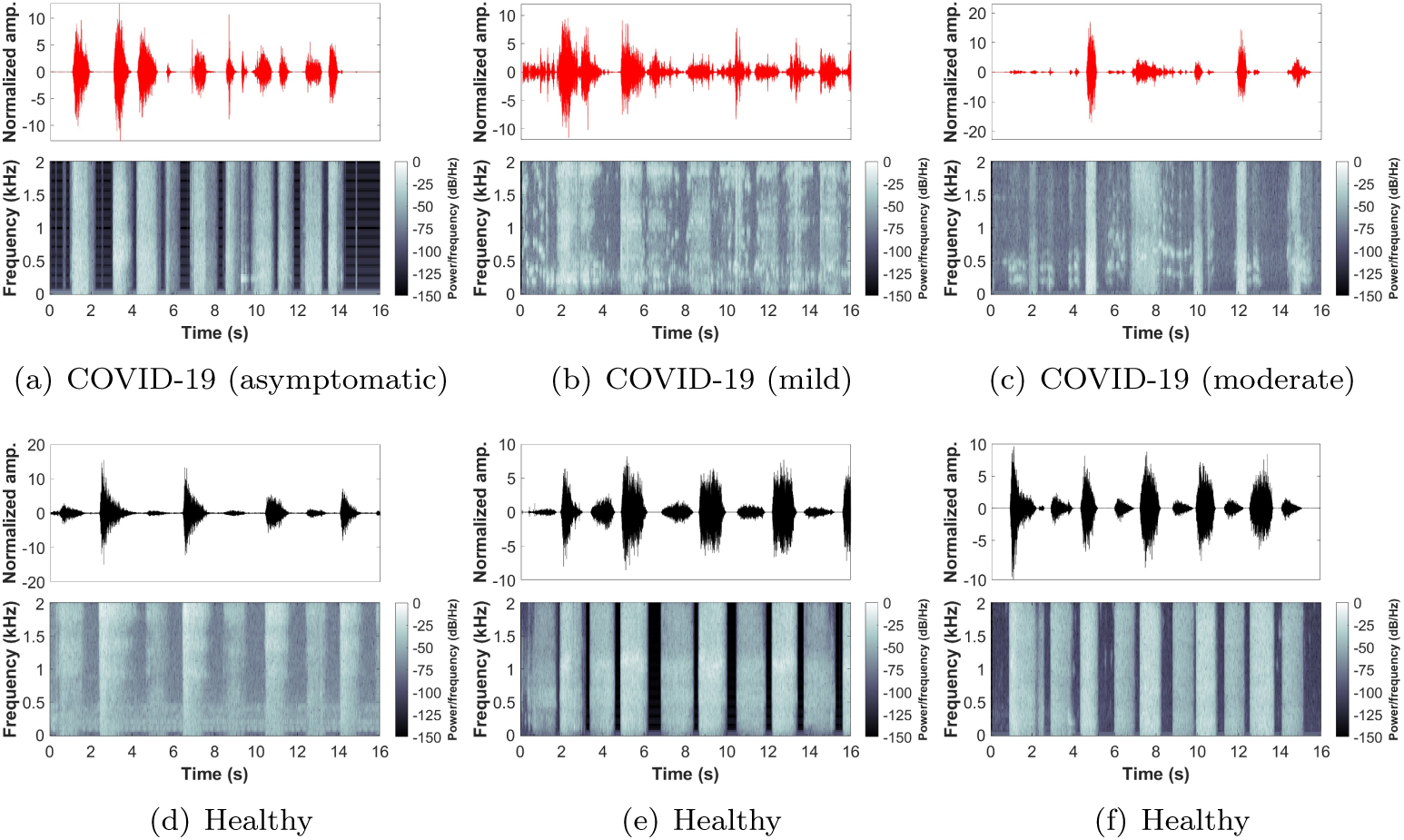
Examples from the shallow breathing sounds recorded via smartphone microphone along with the corresponding spectrogram for: (a-c) COVID-19 subjects (asymptomatic, mild, moderate), (d-f) healthy subjects.

### Deep learning framework

The deep learning framework proposed in this study (Fig. 3) includes a combination of hand-crafted features as well as deep-activated features learned through model’s training and reflected as time-activations of the input. To extract hand-crafted features, various algorithm and functions were used to obtain signal attributes from the original breathing recording and from its corresponding mel-frequency cepstral coefficients (MFCC). In addition, deep-activated learned features were obtained from the original breathing recording through a combined neural network that consists of convolutional and recurrent neural networks. Each part of this framework is briefly described in the following subsections.

**Fig 3.**
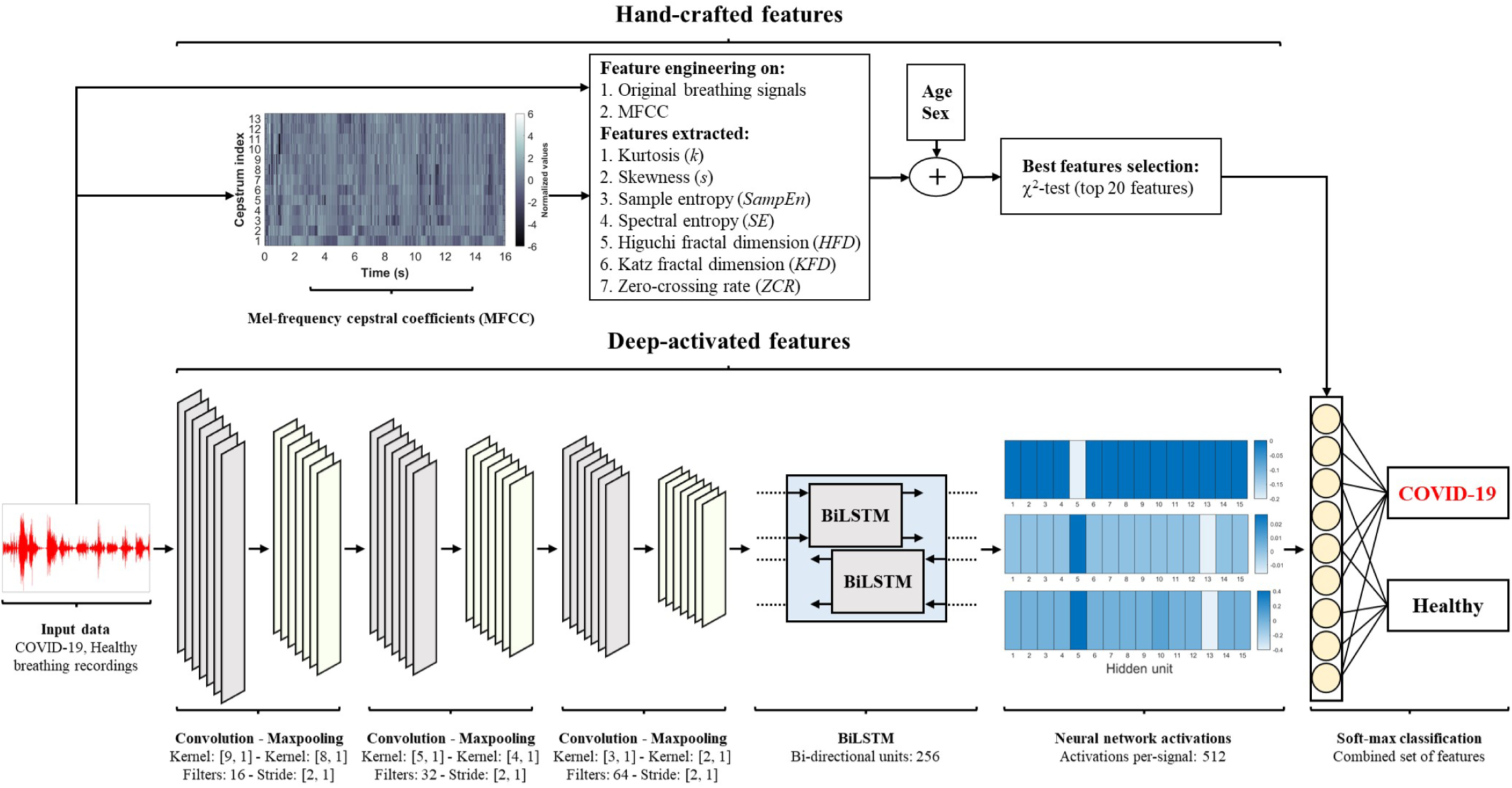
The framework of deep learning followed in this study. The framework includes a combination of hand-crafted features and deep-activated features. Deep features were obtained through a combined convolutional and recurrent neural network (CNN-BiLSTM), and the final classification layer uses both features sets to discriminate between COVID-19 and healthy subjects.

#### Hand-crafted features

These features refer to signal attributes that are extracted manually through various algorithms and functions in a process called feature engineering. The advantage of following such process is that it can extract internal and hidden information within input data, i.e., sounds, and represent it as single or multiple values. Thus, additional knowledge about the input data can be obtained and used for further analysis and evaluation. Hand-crafted features were initially extracted from the original breathing recordings, then, they were also extracted from the MFCC transformation of the signals. The features included in this study are,

##### Kurtosis and Skewness

In statistics, kurtosis is a quantification measure for the degree of extremity included within the tails of a distribution relative to the tails of a normal distribution. The more the distribution is outlier-prone, the higher the kurtosis values, and vice-versa. A kurtosis of 3 indicates that the values follow a normal distribution. On the other hand, skewness is a measure for the asymmetry of the data that deviates it from the mean of the normal distribution. If the skewness is negative, then the data are more spread towards the left side of the mean, while a positive skewness indicates data spreading towards the right side of the mean [27]. A skewness of zero indicates that the values follow a normal distribution. Kurtosis (*k*) and skewness (*s*) can be calculated as,

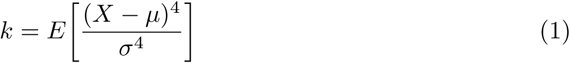

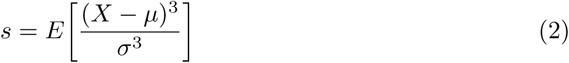

where *X* included input values, *µ* and *σ* are the mean and standard deviation values of the input, respectively, and *E* is an expectation operator.

##### Sample entropy

In physiological signals, the sample entropy (SampEn) provides a measure for complexity contaminated within time sequences. It can be calculated though the negative natural logarithm of a probability that segments of length *m* match their consecutive segments under a value of tolerance (*r*) [28] as follows,

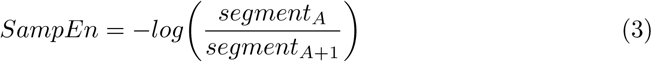

where *segment*_*A*_ is the first segment in the time sequence and *segment*_*A*+1_ is the consecutive segment.

##### Spectral entropy

To measure time series irregularity, spectral entropy (SE) provides a frequency domain entropy measure as a sum of the normalize signal spectral power [29]. Based on Shannon’s entropy, the SE can be calculated as,

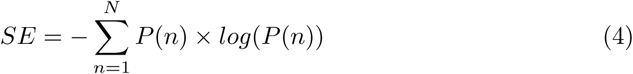

where *N* is the total number of frequency points and *P* (*n*) is the probability distribution of the power spectrum.

##### Fractal dimension

Higuchi and Katz [30, 31] provided two methods to measure statistically the complexity in a time series. More specifically, fractal dimension measures provide an index for characterizing how much a time series is self-similar over some region of space. Higuchi (*HFD*) and Katz (*KFD*) fractal dimensions can be calculated as,

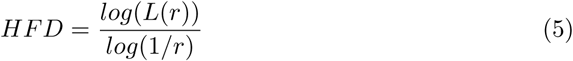

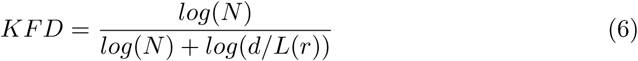

where *L*(*k*) is the length of the fractal curve, *r* is the selected time interval, *N* is the length of the signal, and *d* is the maximum distance between an initial point to other points.

##### Zero-crossing rate

To measure the number of times a signal has passed through the zero point, a zero-crossing rate (ZCR) measure is provided. In other words, ZCR refers to the rate of sign-changes in the signals’ data points. It can be calculated as follows,

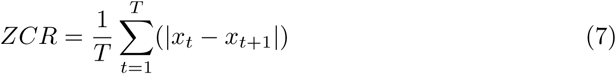

where *x*_*t*_ = 1 if the signal has a positive value at time step *t* and a value of 0 otherwise.

##### Mel-frequency cepstral coefficients (MFCC)

To better represent speech and voice signals, MFCC provides a set of coefficients of the discrete cosine transformed (DCT) logarithm of a signal’s spectrum (mel-frequency cepstrum (MFC)). It is considered as an overall representation of the information contaminated within signals regarding the changes in its different spectrum bands [32, 33]. Briefly, to obtain the coefficients, the signals goes through several steps, namely windowing the signal, applying discrete Fourier transform (DFT), calculating the log energy of the magnitude, transforming the frequencies to the Mel-scale, and applying inverse DCT.

In this work, 13 coefficients (MFCC-1 to MFCC-13) were obtained from each breathing sound signal. For every coefficient, the aforementioned features were extracted and stored as an additional MFCC hand-crafted features alongside the original breathing signals features.

#### 0.0.1 Deep-activated features

These features refer to attributes extracted from signals through a deep learning process and not by manual feature engineering techniques. The utilization of deep learning allows for the acquisition of optimized features extracted through deep convolutional layers about the structural information contaminated within signals. Furthermore, it has the ability to acquire the temporal (time changes) information carried through time sequences [34,35]. Such optimized features can be considered as a complete representation of the input data generated iteratively through an automated learning process. To achieve this, we used an advanced neural network based on a combination of convolutional neural network and bi-directional long short-term memory (CNN-BiLSTM).

##### Neural network architecture

The structure of the network starts by 1D convolutional layers. In deep learning, convolutions refer to a multiple number of dot products applied to 1D signals on pre-defined segments. By applying consecutive convolutions, the network extracts deep attributes (activations) to form an overall feature map for the input data [35]. A single convolution on an input 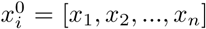, where *n* is the total number of points, is usually calculated as,

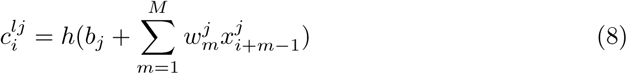

where *l* is the layer index, *h* is the activation function, *b* is the bias of the *j*^*th*^ feature map, *M* is the kernel size, 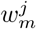 is the weight of the *j*^*th*^ feature map and *m*^*th*^ filter index.

In this work, three convolutional layers were used to form the first stage of the deep neural network. The kernel sizes of each layer are [9, 1], [5, 1], and [3, 1], respectively. Furthermore, the number of filters increases as the network becomes deeper, that is 16, 32, and 64, respectively. Each convolutional layer was followed by a max-pooling layer to reduce the dimensionality as well as the complexity in the model. The max-pooling kernel size decreases as the network gets deeper with a [8, 1], [4, 1], and [2, 1] kernels for the three max-pooling layers, respectively. It is worth noting that each max-pooling layer was followed by a batch normalization (BN) layer to normalize all filters as well as by a rectified linear unit (ReLU) layer to set all values less than zero in the feature map to zero. The complete structure is illustrated in Fig. 3.

The network continues with additional extraction of temporal features through bidirectional LSTM units. In recurrent neural networks, LSTM units allows for the detection of long short-term dependencies between time sequence data points. Thus, it overcomes the issues of exploding and vanishing gradients in chain-like structures during training [34, 36]. An LSTM block includes a collection of gates, namely input (*i*), output (*o*), and forget (*f*) gates. These gates handle the flow of data as well as the processing of the input and output activations within the network’s memory. The information of the main cell (*C*_*t*_) at any instance (*t*) within the block can be calculated as,

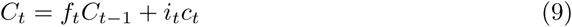

where *c*_*t*_ is the input to the main cell and *C*_*t*−1_ includes the information at the previous time instance.

In addition, the network performs hidden-units (*h*_*t*_) activations on the output and main cell input using a sigmoid function as follows,

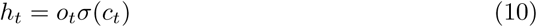

Furthermore, a bi-drectional functionality (BiLSTM) allows the network to process data in both the forward and backward direction as follows,

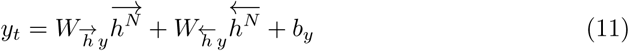

where 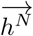 and 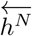 are the outputs of the hidden layers in the forward and backward directions, respectively, for all *N* levels of stack and *b*_*y*_ is a bias vector.

In this work, a BiLSTM hidden units functionality was selected with a total number of hidden units of 256. Thus, the resulting output is a 512 vector (both directions) of the extracted hidden-units of every input.

##### BiLSTM activations

To be able to utilize the parameters that the BiLSTM units have learned, the activations that correspond to each hidden-unit were extracted from the network for each input signal. Recurrent neural network activations of a pre-trained network are vectors that carry the final learned attributes about different time steps within the input [37]. In this work, these activations were the final signal attributes extracted from each input signal. Such attributes are referred to as deep-activated features in this work (Fig. 3). Furthermore, they were concatenated with the hand-crafted features alongside age and sex information and used for the final predictions by the network.

#### 0.0.2 Network configuration and training scheme

Prior to deep learning model training, several data preparation and network fine-tuning steps were followed including data augmentation, best features selection, deciding the training and testing scheme, and network parameters configuration.

##### Data augmentation

Due to the small sample size available, it is critical for deep learning applications to include augmented data. Instead of training the model on the existing dataset only, data augmentation allows for the generation of new modified copies of the original samples. These new copies have similar characteristics of the original data, however, they are slightly adjusted as if they are coming from a new source (subject). Such procedure is essential to expose the deep learning model to more variations in the training data. Thus, making it robust and less biased when attempting to generalize the parameters on new data [38]. Furthermore, it was essential to prevent the model from over-fitting, where the model learns exactly the input data only with a very minimal generalization capabilities for unseen data [39].

In this study, 3,000 samples per class were generated using two 1D data augmentation techniques as follows,

- *Volume control:* Adjusts the strength of signals in decibels (dB) for the generated data [40] with a probability of 0.8 and gain ranging between −5 and 5 dB.
- *Time shift:* Modifies time steps of the signals to illustrate shifting in time for the generated data [41] with a shifting range of [−0.005 to 0.005] seconds.

##### Best features selection

To ensure the inclusion of the most important hand-crafted features within the trained model, a statistical univariate chi-square test (*χ*^2^-test) was applied. In this test, a feature is decided to be important if the observed statistical analysis using this feature matches with the expected one, i.e., label [42]. Furthermore, an important feature indicates that it is considered significant in discriminating between two categories with a *p*-value < 0.05. The lower the *p*-value, the more the feature is dependent on the category label. The importance score can then be calculated as,

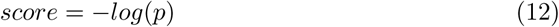

In this work, hand-crafted features extracted from the original breathing signals and from the MFCC alongside the age and sex information were selected for this test. The best 20 features were included in the final best features vector within the final fully-connected layer (along with the deep-activated features) for predictions.

##### Training configuration

To ensure the inclusion of the whole available data, a leave-one-out training and testing scheme was followed. In this scheme, a total of 240 iterations (number of input samples) were applied, where in each iteration, an i^*th*^ subject was used as the testing subject, and the remaining subjects were used for model’s training. This scheme was essential to be followed to provide a prediction for each subject in the dataset.

Furthermore, the network was optimized using adaptive moment estimation (ADAM) solver [43] and with a learning rate of 0.001. The L2-regularization was set to 10^6^ and the mini-batch size to 32.

### Performance evaluation

The performance of the proposed deep learning model in discriminating COVID-19 from healthy subjects was evaluated using traditional evaluation metrics including accuracy, sensitivity, specificity, precision, and F1-score. These metrics can be calculated as,

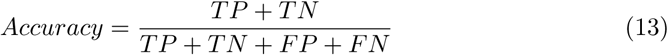

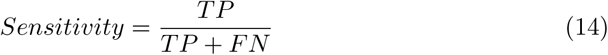

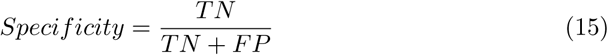

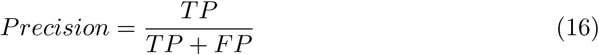

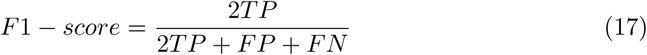

where *TP* is the true positive, *TN* is the true negative, *FP* is the false positive, and *FN* is the false negative numbers in the confusion matrix.

Additionally, the area under the receiver operating characteristic (AUROC) curves was analysed for each category to show the true positive rate (TPR) versus the false positive rate (FPR).

## Results

### Analysis of MFCC

Examples of the 13 MFCC extracted from the original shallow breathing signals are illustrated in Fig. 4 for COVID-19 and healthy subjects. Furthermore, the figure shows MFCC values (after summing all coefficients) distributed as a normal distribution. From the figure, the normal distribution of COVID-19 subjects was slightly skewed to the right side of the mean, while the normal distribution of the healthy subjects was more towards the zero mean, indicating that it better in representing a normal distribution. Tables 2 and 3 show the values of the combined MFCC values, kurtosis, and skewness among all COVID-19 and healthy subjects (mean±std) for the shallow and deep breathing datasets, respectively. In both datasets, the kurtosis and skewness values for COVID-19 subjects were slightly higher than healthy subjects. Furthermore, the average combined MFCC values for COVID-19 were less than those for the healthy subjects. More specifically, in the shallow breathing dataset, a kurtosis and skewness of 4.65±15.97 and 0.59±1.74 was observed for COVID-19 subjects relative to 4.47±20.66 and 00.19±1.75 for healthy subjects. On the other hand, using the deep breathing dataset, COVID-19 subjects had a kurtosis and skewness of 20.82±152.99 and 0.65±4.35 compared to lower values of 3.23±6.06 and −0.36±1.08 for healthy subjects.

**Table 2.** Normal distribution analysis (mean±std) of the combined mel-frequency cepstral coefficients (MFCCs) using the shallow breathing dataset.

| Category |  | Normal distribution analysis |  |  |
| --- | --- | --- | --- | --- |
|  |  | Combined MFCC values | Kurtosis | Skewness |
| COVID-19 | Asymp. | -0.29±0.78 | 1.92±0.94 | 0.45±0.78 |
| | Mild | -0.24±0.70 | 5.46±18.22 | 0.61±1.95 |
| | Moderate | -0.26±0.70 | 2.26±1.85 | 0.60±0.83 |
| | Overall | -0.25±0.72 | 4.65±15.97 | 0.59±1.74 |
| Healthy | | -0.11±0.75 | 4.47±20.66 | 0.19±1.75 |

**Table 3.** Normal distribution analysis (mean±std) of the combined mel-frequency cepstral coefficients (MFCCs) using the deep breathing dataset.

| Category |  | Normal distribution analysis |  |  |
| --- | --- | --- | --- | --- |
|  |  | Combined MFCC values | Kurtosis | Skewness |
| COVID-19 | Asymp. | -0.05±0.69 | 2.61±2.12 | -0.17±0.97 |
| | Mild | -0.15±0.63 | 26.63±15.51 | 0.91±4.95 |
| | Moderate | 0.01±0.58 | 2.54±1.12 | -0.15±0.96 |
| | Overall | -0.12±0.64 | 20.82±12.99 | 0.65±4.35 |
| Healthy | | 0.12±0.60 | 3.23±6.06 | -0.36±1.08 |

**Fig 4.**
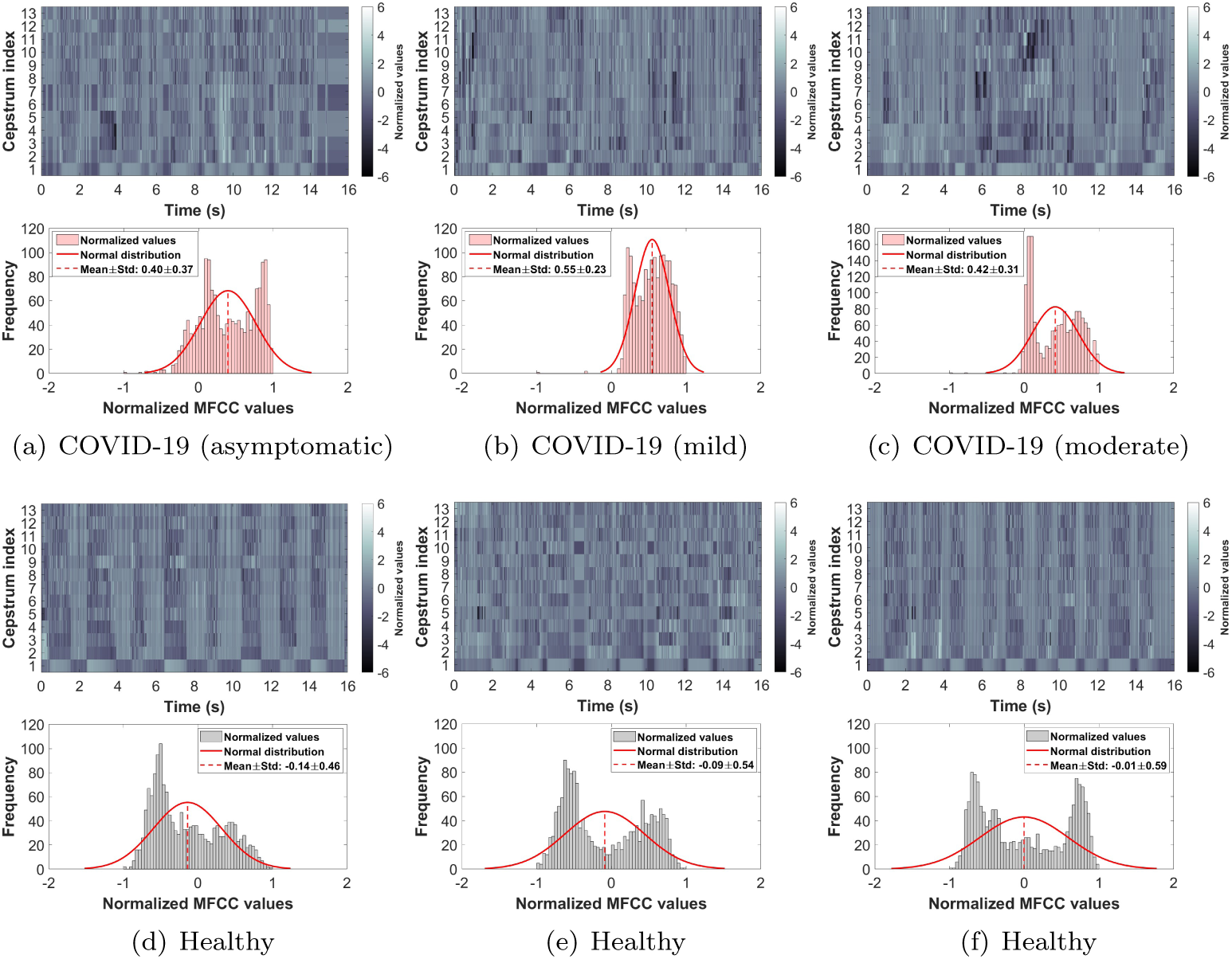
Examples of the mel-frequency cepstral coefficients (MFCC) extracted from the shallow breathing dataset and illustrated as a normal distribution of summed coefficients. (a-c) COVID-19 subjects (asymptomatic, mild, moderate), (d-f) healthy subjects.

### Deep learning performance

The overall performance of the proposed deep learning model is shown in Fig. 5. From the figure, the model correctly predicted 113 and 114 COVID-19 and healthy subjects, respectively, using the shallow breathing dataset out of the 120 total subjects (Fig. 5(a)). In addition, only 7 COVID-19 subjects were miss-classified as healthy, whereas only 6 subjects were wrongly classified as carrying COVID-19. The correct predictions number was slightly lower using the deep breathing dataset with a 109 and 112 for COVID-19 and healthy subjects, respectively. In addition, wrong predictions were also slightly higher with 11 COVID-19 and 8 healthy subjects. Therefore, the confusion matrices show percentages of proportion of 94.20% and 90.80% for COVID-19 subjects using the shallow and deep datasets, respectively. On the other hand, healthy subjects had percentages of 95.00% and 93.30% for both datasets, respectively.

**Fig 5.**
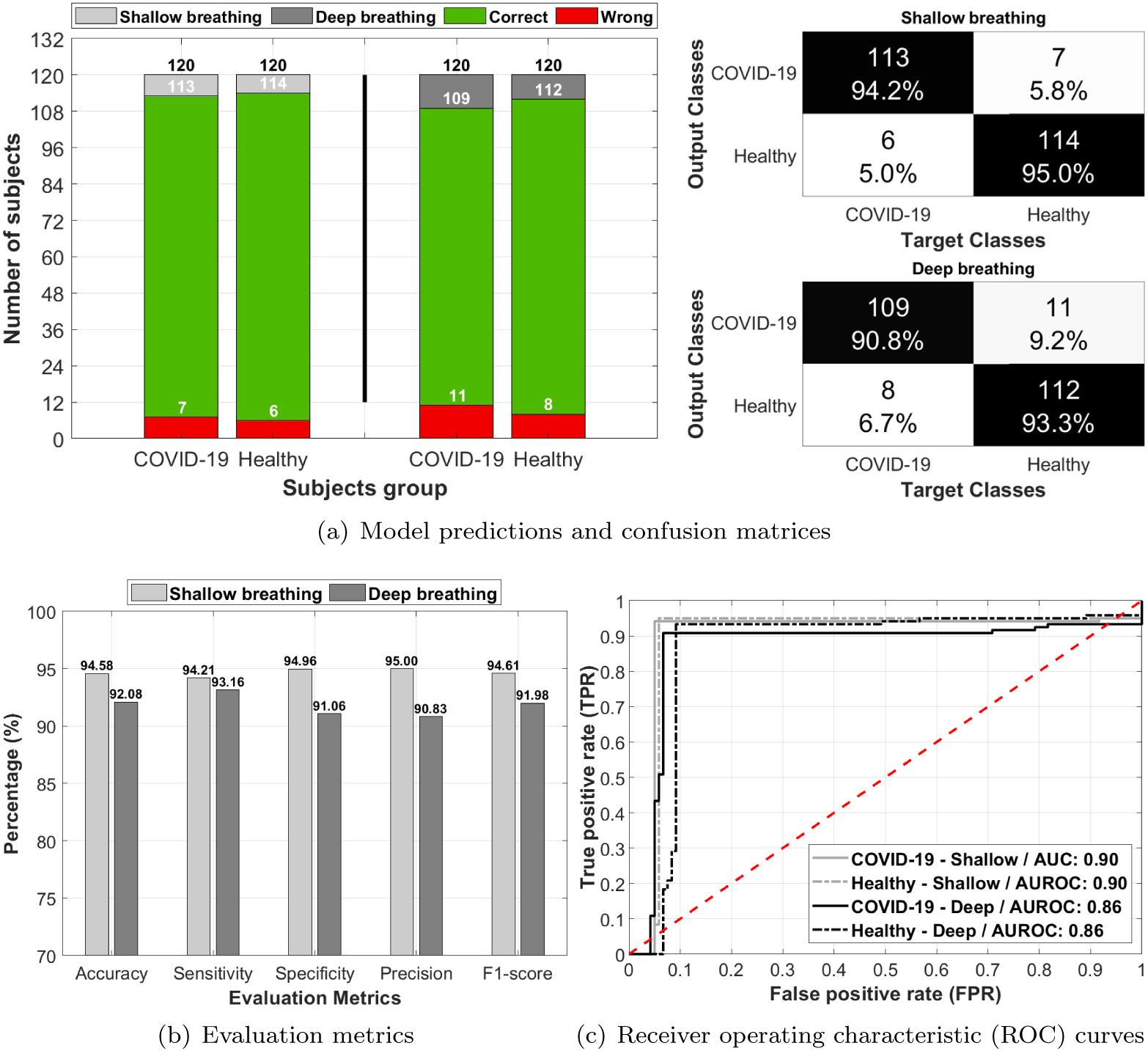
The performance of the deep learning model in predicting COVID-19 and healthy subjects using shallow and deep breathing datasets. (a) model’s predictions for both datasets and he corresponding confusion matrices, (b) evaluation metrics including accuracy, sensitivity, specificity, precision, and F1-score, (d) receiver operating characteristic (ROC) curves and corresponding area under the curve (AUROC) for COVID-19 and healthy subjects using both datasets.

The evaluation metrics (Fig. 5(b)) calculated from these confusion matrices returned an accuracy measure of 94.58% and 92.08% for the shallow and deep datasets, respectively. Furthermore, the model had a sensitivity and specificity measures of 94.21%/94.96% for the shallow dataset and 93.16%/91.06% for the deep dataset. The precision was the highest measure obtained for the shallow dataset (95.00%), where as the deep dataset had the lowest value in the precision with a 90.83%. Lastly, the F1-score measures returned 94.61% and 91.98% for both datasets, respectively.

To analyze the AUROC, Fig. 5(c) shows the ROC curves of predictions using both the shallow and deep datasets. The shallow breathing dataset had an overall AUROC of 0.90 in predicting COVID-19 and healthy subjects, whereas the deep breathing dataset had a 0.86 AUROC, which is slightly lower performance in the prediction process.

Additionally, the model had high accuracy measures in predicting asymptomatic COVID-19 subjects (Fig 6). Using the shallow breathing dataset, the model had a 100.00% accuracy by predicting all subjects correctly. On the other hand, using the deep breathing dataset, the model achieved an accuracy of 88.89% by missing two asymptomatic subjects. It is worth noting that few subjects had close scores (probabilities) to 0.5 using both datasets, however, the model correctly discriminated them from healthy subjects.

**Fig 6.**
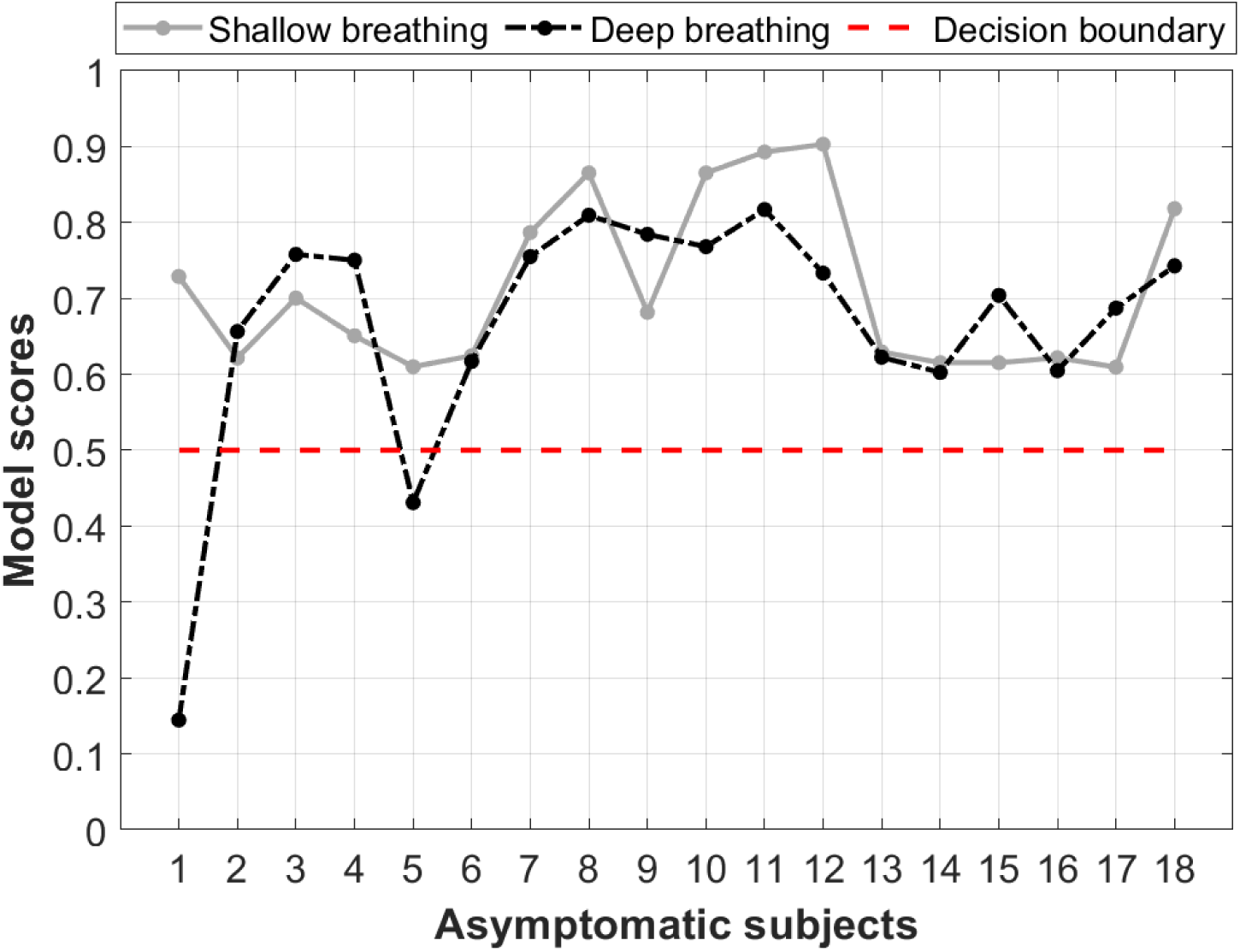
Asymptomatic COVID-19 subjects’ predictions based on the proposed deep learning model. The model had a decision boundary of 0.5 to discriminate between COVID-19 and healthy subjects. The values represent a normalized probability regrading the confidence in predicting these subjects as carrying COVID-19.

## Discussion

This study demonstrated the importance of using deep learning for the detection of COVID-19 subjects, especially those who are asymptomatic. Furthermore, it elaborated on the significance of biological signals, such as breathing sounds, in acquiring useful information about the viral infection. Unlike the conventional lung auscultation techniques, i.e., electronic stethoscopes, to record breathing sounds, the study proposed herein utilized breathing sounds recorded via a smartphone microphone. The observations found in this study (highest accuracy: 94.58%) strongly suggest deep learning as a pre-screening tool for COVID-19 as well as an early detection technique prior to the gold standard RT-PCR assay.

### Smartphone-based breathing recordings

Although current lung auscultation techniques provide high accuracy measures in detecting respiratory diseases [44–46], it requires subjects to be present at hospitals for equipment setup and testing preparation prior to data acquisition. Furthermore, it requires the availability of an experienced person, i.e., clinician or nurse, to take data from patients and store it in a database. Therefore, utilizing a smartphone device to acquire such data allows for a faster data acquisition process from subjects or patients while at the same time, provides highly comparable and acceptable diagnostic performance. In addition, smartphone-based lung auscultation ensures a better social distancing behaviour during lock downs due to pandemics such as COVID-19, thus, it allows for a rapid and time-efficient detection of diseases despite of strong restrictions.

By visually inspecting COVID-19 and healthy subjects’ breathing recordings (Fig. 2), an abnormal nature was usually observed by COVID-19 subjects, while healthy subjects had a more regular pattern during breathing. This could be related to the hidden characteristics of COVID-19 contaminated within lungs and exhibited during lung inhale and exhale [47–49]. Additionally, the MFCC transformation of these recordings (Fig. 4(a-c)) returned similar observations. By quantitatively evaluating these coefficients when combined, COVID-19 subjects had a unique distribution (positively skewed) that can be easily distinguished from the one of healthy subjects. This gives an indication about the importance of further extracting the internal attributes carried not only by the recordings themselves, but rather by the additional MFC transformation of such recordings. Additionally, the asymptomatic subjects had a distribution of values that was close in shape to the distribution of healthy subjects (Fig. 4(a)), however, it was skewed towards the right side of the zero mean. This may be considered as a strong attribute when analyzing COVID-19 patients who do not exhibit any symptoms and thus, discriminating them easily from healthy subjects.

### Diagnosis of COVID-19 using deep learning

It is essential to be able to gain the benefit of the recent advances in AI and computerized algorithms, especially during these hard times of COVID-19 spread worldwide. Deep learning not only provides high levels of performance, it also reduces the dependency on experts, i.e., clinicians and nurses, who are now suffering in handling the pandemic due to the huge and rapidly increasing number of infected patients [50–52]. Recently, the detection of COVID-19 using deep learning has reached high levels of accuracy through two-dimensional (2D) lung CT images [53–55]. Despite of such performance in discriminating and detecting COVID-19 subjects, CT imaging is considered high in cost and requires extra time to acquire testing data and results. Furthermore, it utilizes excessive amount of ionizing radiations (X-ray) that are usually harmful to the human body, especially for severely affected lungs. Therefore, the integration of biological sounds, as in breathing recordings, within a deep learning framework overcomes the aforementioned limitations, while at the same time provides acceptable levels of performance.

The proposed deep learning framework had high levels of accuracy (94.58%) in discriminating between COVID-19 and healthy subjects. The structure of the framework was built to ensure a simple architecture, while at the same time to provide advanced features extraction and learning mechanisms. The combination between hand-crafted features and deep-activated features allowed for maximized performance capabilities within the model, as it learns through hidden and internal attributes as well as deep structural and temporal characteristics of recordings. The high sensitivity and specificity measures (94.21% and 94.96%, respectively) obtained in this study prove the efficiency of deep learning in distinguishing COVID-19 subjects (AUROC: 0.90). Additionally, it supports the field of deep learning research on the use of respiratory signals for COVID-19 diagnostics [21, 56]. Alongside the high performance levels, it was interesting to observe a 100.00% accuracy in predicting asymptomatic COVID-19 subjects. This could enhance the detection of this viral infection at a very early stage and thus, preventing it from developing to mild and moderate conditions or spreading to other people.

Furthermore, this high performance levels were achieved through 1D signals instead of 2D images, which allowed the model to be simple and not memory exhausting. In addition, due to its simplicity and effective performance, it can be easily embedded within smartphone applications and internet-of-things tools to allow real-time and direct connectivity between the subject and family for care or healthcare authorities for services.

### Clinical relevance

The utilization of smartphone-based breathing recordings within a deep learning framework may have the potential to provide a non-invasive, zero-cost, rapid pre-screening tool for COVID-19 in low-infected as well as servery-infected countries. Furthermore, it may be useful for countries who are not able of providing the RT-PCR test to everyone due to healthcare, economic, and political difficulties. Furthermore, instead of performing RT-PCR tests on daily or weekly basis, the proposed framework allows for easier, cost effective, and faster large-scale detection, especially for counties/areas who are putting high expenses on such tests due to logistical complications. Alongside the rapid nature of this approach, many healthcare service could be revived significantly by decreasing the demand on clinicians or nurses. In addition, due to the ability of successfully detecting asymptomatic subjects, it can decrease the need for extra equipment and costs associated with further medication after the development of the viral infection in patients.

Clinically, it is better to have a faster connection between COVID-19 subjects and medical practitioners or health authorities to ensure continues monitoring for such cases and at the same time maintain successful contact tracing and social distancing. By embedding such approach within a smartphone applications or cloud-based networks, monitoring subjects, including those who are healthy or suspected to be carrying the virus, does not require the presence at clinics or testing points. Instead, it can be performed real-time through a direct connectivity with a medical practitioners. In addition, it can be completely done by the subject himself to self-test his condition prior to taking further steps towards the RT-PCR assay. Therefore, such approach could set an early alert to people, especially those who interacted with COVID-19 subjects or are asymptomatic, to go and further diagnose their case. Considering such mechanism in detecting COVID-19 could provide a better and well-organized approach that results in less demand for clinics and medical tests, and thus, enhances back the healthcare and economic sectors in various countries worldwide.

## Conclusion

This study suggests smartphone-based breathing sounds as a promising indicator for COVID-19 cases. It further recommends the utilization of deep learning as a pre-screening tool for such cases prior to the gold standard RT-PCR tests. The overall performance found in this study (accuracy 94.58%) in discriminating between COVID-19 and healthy subjects shows the potential of such approach. This study paves the way towards implementing deep learning in COVID-19 diagnostics by suggesting it as a rapid, time-efficient, and no-cost technique that does not violate social distancing restrictions during pandemics such as COVID-19.

## Data Availability

Data are publicly available at Coswara Project

https://github.com/iiscleap/Coswara-Data

## Acknowledgement

This work was supported by a grant (award number: 8474000132) from the Healthcare Engineering Innovation Center (HEIC) at Khalifa University, Abu Dhabi, UAE, and by grant (award number: 29934) from the Department of Education and Knowledge (ADEK), Abu Dhabi, UAE.

